# Impact of Superspreaders on dissemination and mitigation of COVID-19

**DOI:** 10.1101/2020.05.17.20104745

**Authors:** Kim Sneppen, Robert J. Taylor, Lone Simonsen

## Abstract

**Background:** The draconian measures used to control COVID-19 dissemination have been highly effective but only at enormous socioeconomic cost. Evidence suggests that “superspreaders” who transmit the virus to a large number of people, play a substantial role in transmission; recent estimates suggest that about 1-20% of people with the virus are the source for about 80% of infections. We used an agent-based model to explore the interplay between social structure, mitigation and superspreading.

**Methods:** We developed an agent-based model with a subset of “superspreader” agents that transmit disease far more efficiently. These agents act in a social network that allows transmission during contacts in three sectors: “home,” “work/school” and “other”. We simulated the effect of various mitigation strategies that limit contacts in each of these sectors, and used the model to fit COVID-19 mortality data from Sweden.

**Findings:** Reducing contacts in the “other” sector had a far greater impact on epidemic trajectory than did reducing “home” or “work/school” contacts; this effect was substantially enhanced when the infectivity of children was reduced relative to that of adults. The model fit Swedish hospitalization data with reasonable assumptions about the effect of Sweden’s mitigation policies on contacts in the different sectors.

**Interpretation:** Our results suggest COVID-19 could be controlled by limiting large gatherings and other opportunities for contacts between people in restaurants, sporting events, concerts and worship services) while still allowing regular contacts in the home or at work and school.

Research in context

Evidence before this study
Superspreading events have long been known to be important in the epidemiology of many infectious diseases, including tuberculosis, measles, Ebola and SARS. Since the emergence of SARS-CoV-2, epidemiologic analyses have inferred substantial individual-level variation in transmissibility, with an estimated 1% to 20% of infected persons causing about 80% of all COVID-19 cases.

Added value of this study
We developed an agent-based socially structured model to simulate the effect of superspreaders in COVID-19 transmission in the context of country-wide “lockdown” policies. These simulations indicate that COVID-19 can be effectively mitigated by limiting contacts between people who otherwise rarely meet, while allowing home and most work/school contacts to continue.

Implications of all available evidence
It is crucial to include heterogeneity in individual infectiousness when modeling the impact of mitigation strategies on observed COVID-19 epidemic patterns. Reducing opportunities for superspreading by limiting random contacts outside home and work could be the most effective way to control COVID-19. Our findings suggest why the epidemic has continued to decline following re-opening of work and school in European countries. The superspreader phenomenon may also explain the variability in COVID-19 incidence in rural and urban areas within a country.

## Introduction

Countries worldwide have responded to the COVID-19 pandemic by implementing an unprecedented lock-down strategy: banning large gatherings of any kind, closing schools and most workplaces, and prohibiting many normal activities such as walking in parks, eating at restaurants and attending large gatherings such as sporting events and religious services. These lock-downs have dramatically limited disease transmission, but only at enormous socioeconomic cost; however, Sweden became an outlier among European countries by only prohibiting large gatherings and relying on the population to practice social-distancing while leaving schools and workplaces open.^1^ Unfortunately, we do not know enough about which aspects of such lockdowns are most effective. Given that many countries are already in the process of easing lockdown restrictions, the relative contribution of reducing contacts at home, schools, workplaces and other sectors of society needs to be thoroughly understood as soon as possible.

“Superspreaders”—single individuals who infect a large number of people in a short time—must be considered when trying to determine which mitigation strategies work best. Heterogeneity in transmission risk is a well known phenomenon in infectious diseases^2,3^. Superspreaders had a large impact on the transmission dynamics of the recent coronavirus threats SARS^4^ and MERS^5^. In 2005, Lloyd-Smith et al. concluded that ‘superspreading events’ are important in epidemics of many infectious diseases, with about 20% of the infected population being responsible for about 80% of transmission events^6^.

Considerable evidence indicates that superspreaders are also important in the spread of COVID-19^2^, and a list of 1100 outbreaks around the world involving superspreaders have been compiled^7^. Examples include an outbreak in South Korea in which a single infected person at a night club causing at least 50 new infections, and a 2.5 hour choir rehearsal in Skagit, Washington where 52 out of 61 attendees were infected.^8^ Such events, as well as outbreaks in prisons and hospitals, are reminiscent of documented superspreader events in the 2003 SARS-CoV outbreak. Morever, multiple studies of COVID-19 have quantitatively assessed the heterogeneity of infectivity among infected individuals, finding that 1% to 20% of infected people cause about 80% of new infections.^9–13^

Given the evidence that superspreaders are important in COVID-19 transmission, models should not rely on a single parameter such as the basic reproductive number (*R*_0_), because doing so obscures the considerable impact of individual variation in infectivity on an epidemic’s trajectory.^3,15–17^ Agent based models, however, are very well-suited to investigate the role of superspreaders. Like standard compartmental SEIR models, they can easily reproduce the epidemic curves observed in a population. Unlike purely compartmental models, however, agent-based models can adjust individual infectivity and mimic repeated social interactions within defined groups. In an agent-based model an agent goes to the same workplace in the morning and home to the same household at night. In contrast, inhabitants of standard compartmental models go to a new workplace and home to a new family in every time step. Agent-based models thereby can capture effect the disease saturating a household or workplace as the available susceptible agents become infected. These are exactly the properties needed to investigate how superspreaders might affect the various mitigation strategies government officials might choose to combat COVID-19.

We have used agent-based models to investigate how superspreaders in a population might affect non-pharmaceutical mitigation efforts to control COVID-19. Our findings suggest that COVID-19 might have an Achilles Heel, namely that limiting contacts in the portion of the social environment where many new and random contacts are encountered and in which superspreading events can occur likely has a far greater benefit than closing workplaces, schools and other places where repeated contacts occur among small social circles.

## Methods

We developed an age-stratified, agent-based model with three simulated sectors of social contact through which the disease can be transmitted. Each agent was assigned to one “home” and one “work/school” unit and participated in random “other” contacts. Agents were stratified in 10 year intervals and assigned age-dependent social activity ^18^. Each home had an average of 2.1 members, in which adults were in the same or adjacent age band and children were 20-40 years younger than their parents. Adult agents 20-70 years of age were assigned a “workplace,” a Poisson distributed cluster of average size 6 agents; to simulate interactions between workplaces, each agent’s connections were assigned to two random persons outside this cluster. Agents under 20 years old were assigned a “school” class of 18 members; each school class was also assigned two “teachers” age 20-70, which constituted the teachers’ workplace. Agents older than 70 years were not assigned to a workplace. “Other” contacts were chosen at random from the entire population.

Progression of disease was modeled in a Susceptible, Exposed, Infected, Recovered (SEIR) framework, with agents passing through each stage according to preset rules (Figure 1). The exposed period was set to 5 days, extending from infection to symptom onset. Agents became infectious 2.5 days after infection and remain so through day 3 after symptom onset^19^. All transitions between stages were implemented as a corresponding probability per time to pass to next stage. Age-dependent conditional probabilities governed progression from symptomatic illness to hospitalization and intensive care (ICU) use (Table S1)^20^; probability of death was calibrated to produce a rate of 0.3%.

**Figure 1:**
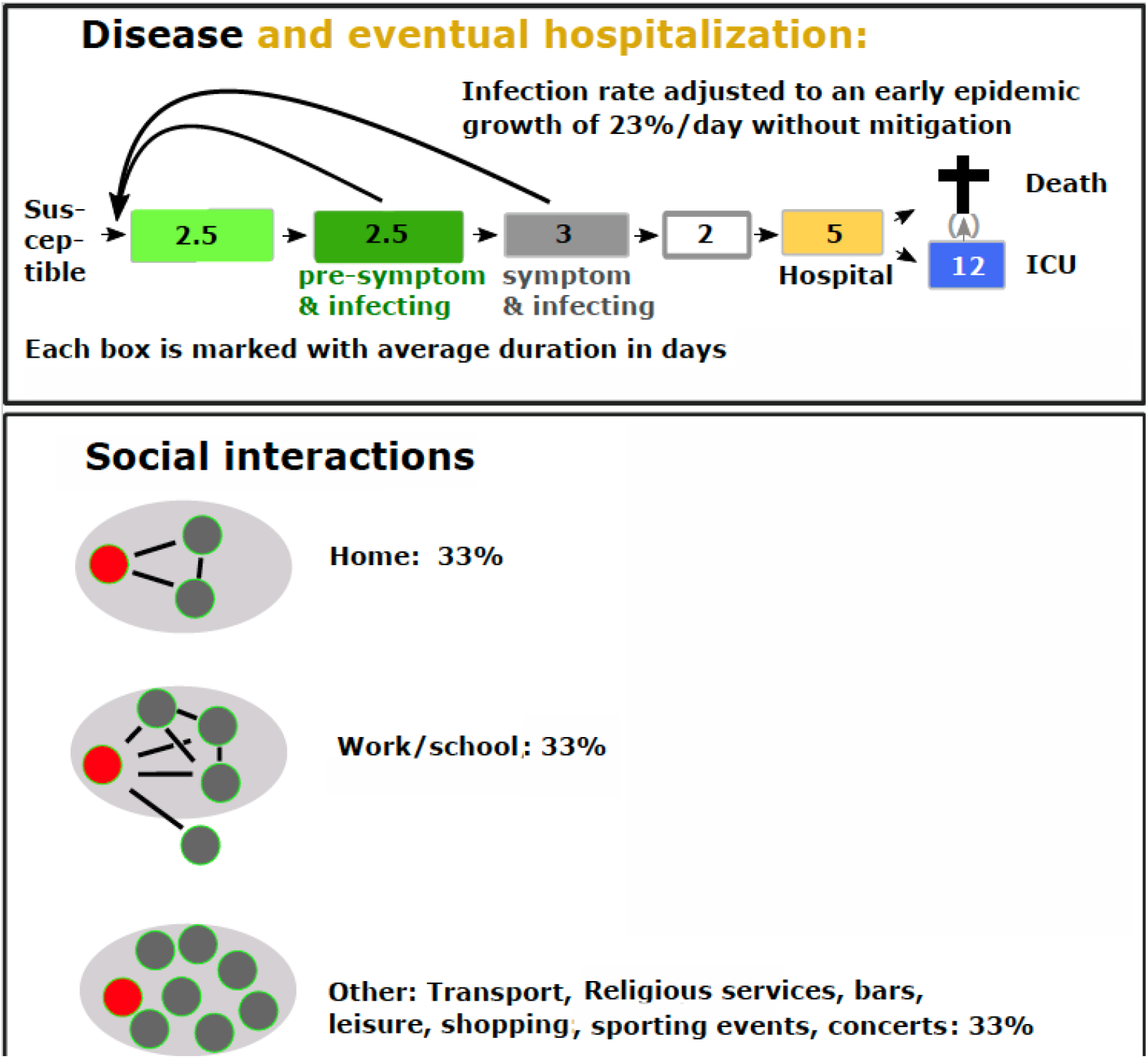
Schematic representation of our agent-based model. Agents progress in an SEIR framework, becoming infectious 2.5 days before symptom onset [12] (panel A). Most agents enter the recovered pool after 5 days with symptoms; some are hospitalized, and some of those enter the ICU or die according to conditional probabilities given in Table S1. The model’s social network (panel B) allows contacts in three sectors, “Home”, “Work/School” and “Other”, with equal frequency.

**Figure 2:**
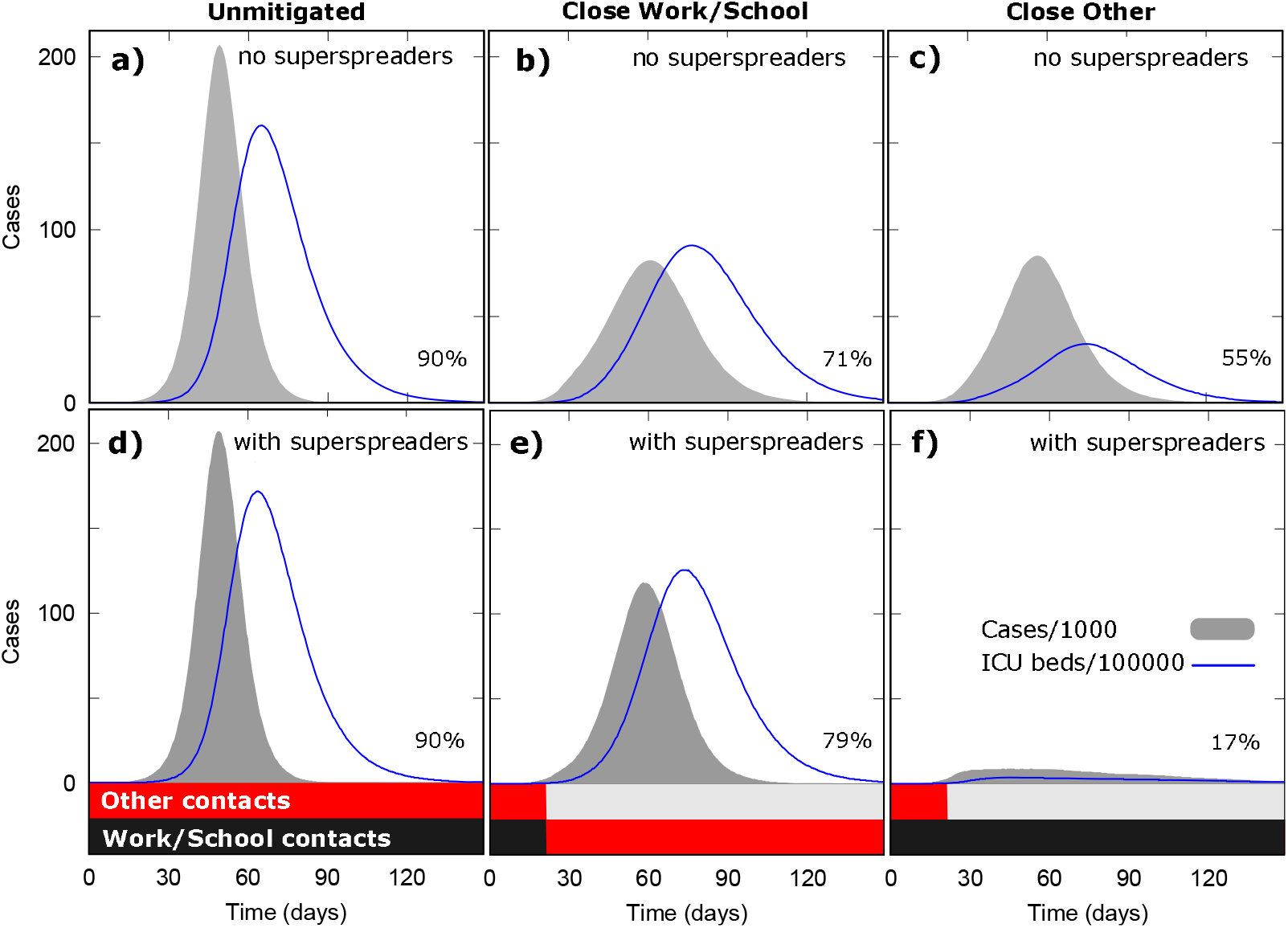
The impact of mitigation on modeled incidence and ICU use. The number of symptomatic agents per 1000 (gray) and ICU cases per 100,000 (blue line) is scarcely affected by the presence of superspreaders (10% of population, s_i_=50) when no mitigation is applied **(panels a, d)**, and only modestly more so when “work/school” contacts are eliminated **(panels b, e)**. However, eliminating “other” contacts **(panels c, f)** has a far greater impact on epidemic trajectory when superspreaders are included. The black and red stacked bar chart shows the proportion of “work/school” and “other” contacts allowed.

Most agents were assigned an infection activity parameter (*s*_*i*_) of 1, indicating that the agent has one chance of transmitting the virus at a given contact. A chosen proportion of agents were designated as superspreaders, with *s*_*i*_ >1; unless otherwise noted, 10% of agents were assigned *s*_*i*_ = 50. Simulations were run in a population of 1 million seeded with 100 infected agents.. In defined time steps Δt within an agent’s infectious period, each infected agent was chosen for a contact with an age-dependent probability. For each chosen agent we assigned a contact in one of the three sectors: home, work/school, other. The were selected with probabilities such that they occur in a ratio of 1:1:1 across the population^18^ Contacts were selected contacts so that 1/3 occurred in each of our three social sectors, after Mossong et al. (2008), which weighted the “home” sector at 19% to 50% of all contacts, the “work/school” sector at 23% to 37% and the remaining sectors at 27% to 44%.^18^ Potential targets for infection were also selected based on the age dependent probability.

At each contact, the disease was transmitted with probability P_t_ = β s_i_ Δt, where the rate constant β is adjusted to fit the observed 23% growth per day of an unmitigated COVID-19 epidemic^21^. The time step length was chosen to ensure that the probability of infection was always less than 1. We simulated mitigation strategies by not permitting infection during a portion of contacts in one or more of the contact categories. We began mitigation when the infected population reached 1% of the total.

In a sensitivity analysis we explored the effect of agents having a gamma-distributed infectivity β *s*_*i*_, where *s*_*i*_ was drawn from a Gamma distribution P(*s*) proportional to *s^{k-1}* exp(-*k* s) with continuous s>0 and where *k* is the dispersion parameter.^6^ For each simulation the rate β is adjusted to fit the exponential growth of an unmitigated epidemic of 23% per day. Attempts at infection were implemented as described above, using discrete time steps Δt in which agent *i* has a probability to transmit P_t_ = β *s*_*i*_ Δt.

We fitted model-generated epidemic curves to real-life daily numbers of COVID-19 fatalities in Sweden.^22^ To mimic Sweden’s unusual mitigation strategy, we left “work” contacts at 100% and varied 1) the date of onset of the mitigation effort, 2) the assumed proportion of cases at the start of mitigation and 3) the proportion of “other” contacts left active to achieve best chi-squared fit to the data. The fit assumes an overall mortality rate of 0.3% for COVID-19 in Sweden; if this mortality were higher, then the the fitted number of infected persons at onset of mitigation would be smaller.

The funders of the study had no role in the study design, data collection, data interpretation, or writing of this paper. The corresponding author had full access to all the data in the study and had final responsibility for the decision to submit for publication.

## Results

We first simulated epidemic trajectories in our socially structured model both without and with superspreaders. We began with all contacts allowed, then applied two simulated mitigation strategies, removing in turn “work/school” and “other” contacts (Figure 3). As expected, when we allowed all contacts (i.e., no mitigation), the presence of superspreaders changed the epidemic trajectory very little compared to the model with no superspreaders (panels a, d). When “work/school” contacts were eliminated, the epidemic curves with and without superspreaders were still similar: the epidemics had been broadened and flattened somewhat, with slightly lower peaks in cases and in ICU demand (panels b, e). However, including superspreaders in the model greatly increased the impact of preventing “other” contacts (panels c, f) compared to the cases where superspreaders were not present. The projected number of cases and ICU admissions were both substantially smaller when superspreaders were included in the model, and peak ICU demand was far smaller. In terms of cumulative infections, with no superspreaders total cases dropped from 89% of all agents in the unmitigated case to 79% and 55% of the simulated population when closing the “work/school” and “other” sectors, respectively. But with superspreaders, closing “other” contacts had a dramatic impact, essentially halting the epidemic in its tracks and reducing cumulative cases to only 17% of the population, with an even greater impact on ICU demand.

**Figure 3.**
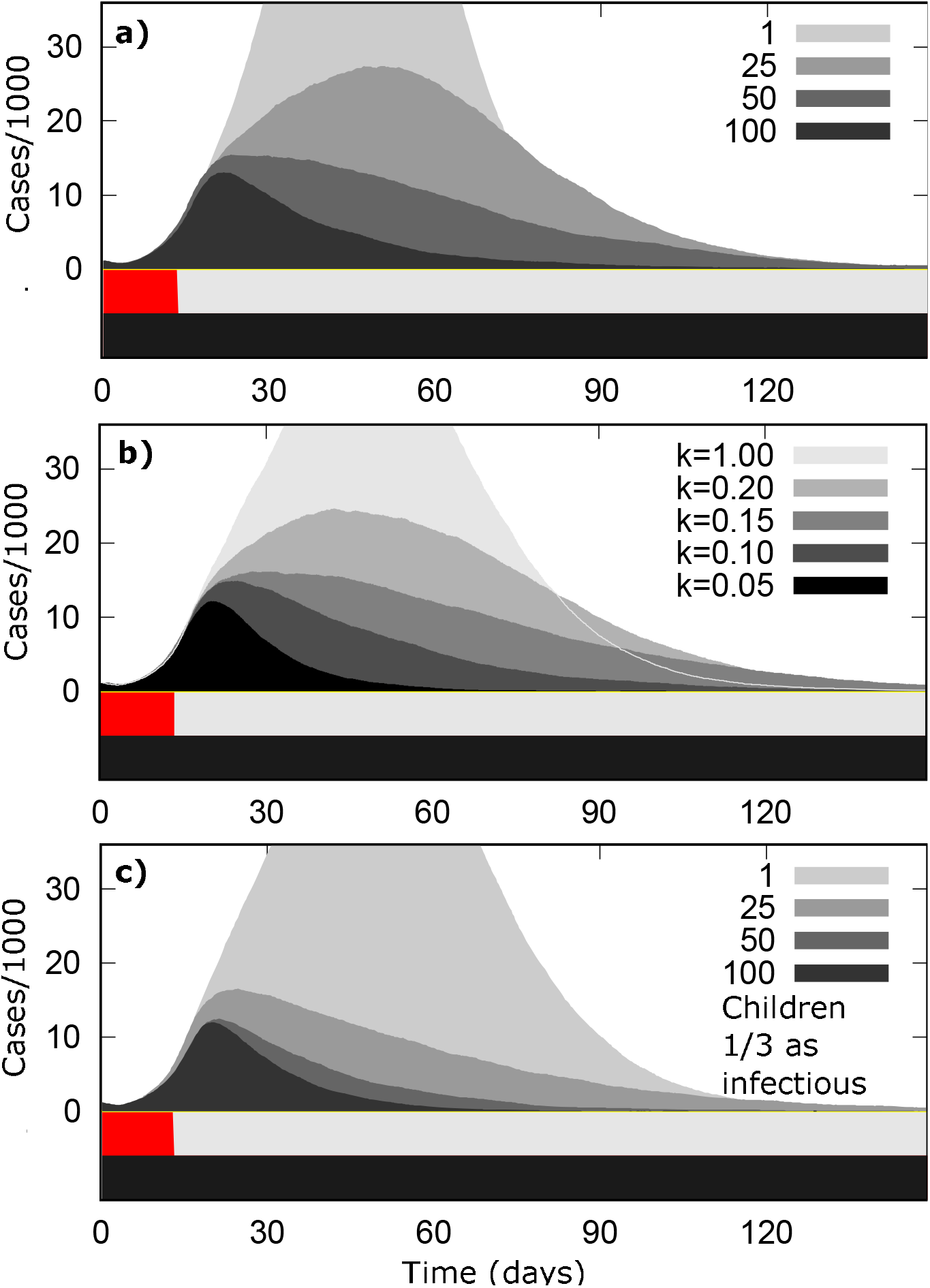
Sensitivity of model results to changing superspreader strength si (a), distributing infectiousness on a gamma distribution with various dispersion factor *k* (b) and reducing infectivity of children (c). In all cases, the probability of transmission was adjusted to produce an initial growth rate of 23%/day, and mitigation was instituted when infected agents reached 1% of the total population. The black and red stacked bar chart shows the proportion of “work/school” and “other” contacts allowed.

Next, we assessed the sensitivity of the impact of reducing “other” contacts in the superspreader model in several ways. First, we varied the infectivity we assigned to designated superspreaders (*s*_*i*_,) from the base model value of 50 (Figure 3a). As expected higher values for *s*_*i*_, increased the impact of closing “other” contacts and vice versa. We also increased from zero the proportion of “other” contacts allowed (Figure S1). Second, we assigned every agent a value for *s*_*i*_ according to a gamma distribution, then varied the dispersion factor k in the distribution (Figure 3b). A *k* of about 0.13 produced an impact similar to an *s*_*i*_, of 50 with 10% of the population is designated as a superspreader. Third, we increased the size of the workplace from an average of 6 members to 12 (Figure S2b). We also varied the ratio of contacts from 33%:33%:33% to 40%:40%:20% for “home,” “work/school”, and “other” sectors (Figure S2a).

Because considerable confusion surrounds the contribution of children in the dissemination of COVID-19, we also conducted a sensitivity analysis in which children contributed less than adults to the epidemic.^23^ We reduced *s*_*i*_ for agents assigned an age of <20 years by a factor of three, i.e., to 0.33 for most and to 16.67 for superspreaders; we also reduced both the infectivity and infection rate (i.e. “exposure”) of children by half. In both cases, a smaller role for children in transmission increased the benefit of reducing “other” contacts (Figures 4c, S3).

**Figure 4.**
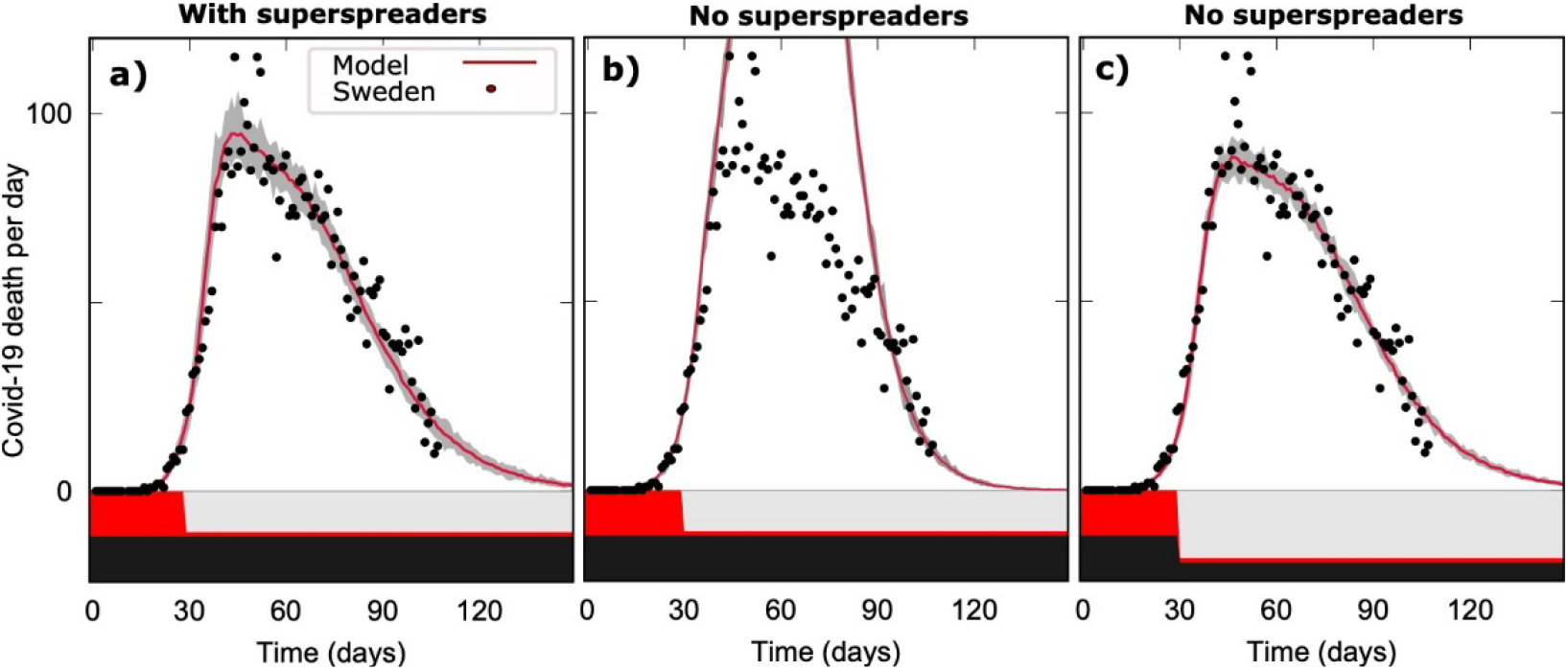
Model fit to daily COVID-19 fatalities in Sweden. Red lines shows average of 20 model runs; gray area is area into which 95% of trajectories fall. Best fit occurred when “other” contacts were reduced to 90% of normal (panel a). However, when superspreaders were removed from the model, those parameters no longer fit (panel b). Fit could be restored if “work/school” contacts were reduced to 40% of normal to achieve best fit while still reducing “other” contacts by 90%. The black and red stacked bar chart shows the proportion of “work/school” and “other” contacts allowed.

Last, we tested the model’s ability to simulate real-life mortality data from Sweden’s experience with COVID-19 up to June 7 2020. To mimic Sweden’s relaxed COVID-19 policy, we fixed work/school contacts at 100%. Best fit occurred when “other” contacts were reduced by 90% (Figure 4a). As might be expected, when we removed superspreaders from the model but used the same parameters, the generated curve no longer fit the data; however, reducing the “work/school” contacts by 60%, while maintaining the 90% reduction in “other” contacts, restored a good fit to the data.

## Discussion

Policy makers face excruciating choices as they seek to open up economies and societies as much as possible without causing a surge in COVID-19 cases that would kill many and overwhelm health care systems, especially by exceeding available ICU beds and mechanical ventilators needed to keep critically ill COVID-19 patients alive. Our model results suggest that the to the extent that superspreaders are driving the epidemic, those choices should tend toward allowing workplaces, schools and other groups with a relatively small number of regular participants to continue, while activities that bring together people who otherwise would not come into contact, as occurs at sporting events, restaurants and bars, large parties and worship services. To our knowledge, this is the first agent-based modeling study to test how superspreaders change the impact of different mitigation strategies on an epidemic.

Evidence is mounting that superspreaders play a large role in COVID-19 transmission. Studies based on observational data estimate that 1% to 20% of infected cases cause ∼80% new transmissions. This means that the majority of cases cause less than one secondary case and thus cannot sustain the epidemic on their own. The attack rate in households is in fact low,^10^ as documented and among Chinese household contacts (15%)^10^ and in the context of a superspreading event in a South Korean call center (16%).^24^

We found that the large benefit from closing “other” contacts was robust in all of our sensitivity analyses. The impact of closing “other” contacts changed under these various conditions, but the effect was always substantially greater than when superspreaders were not in the model. In particular, regardless of how we reduced the role of children in the spread of the virus, we got an even stronger effect of mitigating of “other” sector contacts (Figure 3 and S3

We further tested the model by fitting it to daily numbers of Swedish COVID-19 deaths, and found that it could be fit to real-world data. Sweden famously did not close its schools and workplaces, instead deciding to increase overall social distancing in public places and to prohibit large events^25^. The parameters that produced the best fit when superspreaders were included in the model broadly reflected what actually occurred in Sweden—workplaces and schools were not closed, but other contacts were limited. When we removed superspreaders those parameters no longer produced a curve that fit the data. We restored the fit, but only by removing a substantial portion of “work/school” contacts as well as the 90% of the other contacts needed to fit the “with superspreaders” model. This is not realistic, as work and especially schools remained open in Sweden throughout.

Our model demonstrates that the presence of superspreaders substantially favors mitigating “other” sector contacts over mitigating the home and work/school sectors. When the “other” sector is closed, “work/school” contacts constitute the majority of remaining individual contacts. But because the number of connections is limited in a “work/school” social unit, a superspreader soon infects all the available susceptible contacts, and that saturation limits the potential to transmit widely. When the “other” sector is open, however, the superspreader faces no such limitation. This means that an epidemic driven by superspreaders is fueled by the *diversity* of personal contacts, and is less dependent on the *duration* of contacts. Therefore closing the “work/school” sector in the model provides less benefit than closing the “other” sector.

Our model results indicate that mitigation policies designed to limit transmission during random contacts between people not otherwise linked will likely provide a far greater benefit than closing workplaces and schools. That is, mitigation strategies should aim to limit the opportunities for large numbers of people to come into contact with a superspreader. These opportunities include large events as well as contacts in other public spaces such as public transportation. For situations where such contacts cannot be avoided, steps such as wearing face masks and moving events outdoors could have the an impact similar to that of preventing the contact altogether.

Our study is subject to several limitations. The most obvious is a model’s simplicity compared to the complex reality of human society. We relegated all random (non-repeating) contacts to the “other” category, so that contacts with known persons occurred only through fixed social networks at home or the workplace. In reality, many interactions in the “other” sector would be with familiar persons such as friends and extended family, while some of the interactions in the “work/school” sector would involve unknown persons, such as in cafeterias, conferences and workshops. But even though the reality of where these random contacts take place unquestionably differs from our model, the sensitivity analyses indicate that does not undercut end result.

We chose to create “superspreaders” in the model by increasing the infectivity of a fixed minority of those infected, as if it were a biological property such as high viral load. But many factors are needed to create a super-spreading event. These may include specific behaviors of infected persons such as coughing, singing, shouting, as well as being in a crowded situation with many random contacts. However, regardless of biological mechanism, superspreading will be largely preventable with a policy that limits opportunity to make many contacts.

In closing, we note that our results may help to explain observed differences in patterns of COVID-19 transmission in different parts of a single country, especially between urban and rural areas. The amount of time people spend with other people is likely similar in the city and countryside, but the diversity in contacts would be far higher in urban areas, thereby allowing superspreaders to transmit the virus to their full potential. Superspreading also likely explains the surprising success of country lock-downs in Europe, as well as the lack of a resurgence following reopening of the work and school sector in Denmark and elsewhere. Superspreading may indeed be the Achilles Heel of the novel coronavirus that allows it to be at least partially controlled at a bearable socio-economic cost.

## Data Availability

No data.

## Funding

KS received funding from the EU Horizon 2020 research program under ERC grant No. 740704, and LS and RJT from the Carlsberg Foundation in Denmark.

## Acknowledgements

We thank Raul Donangelo, Viggo Andreasen, Andreas Eilersen and Bjarke Frost Nielsen for enlightening discussions and multiple corrections and suggestions to the manuscript.

## Supplementary Material

**Figure S1.**
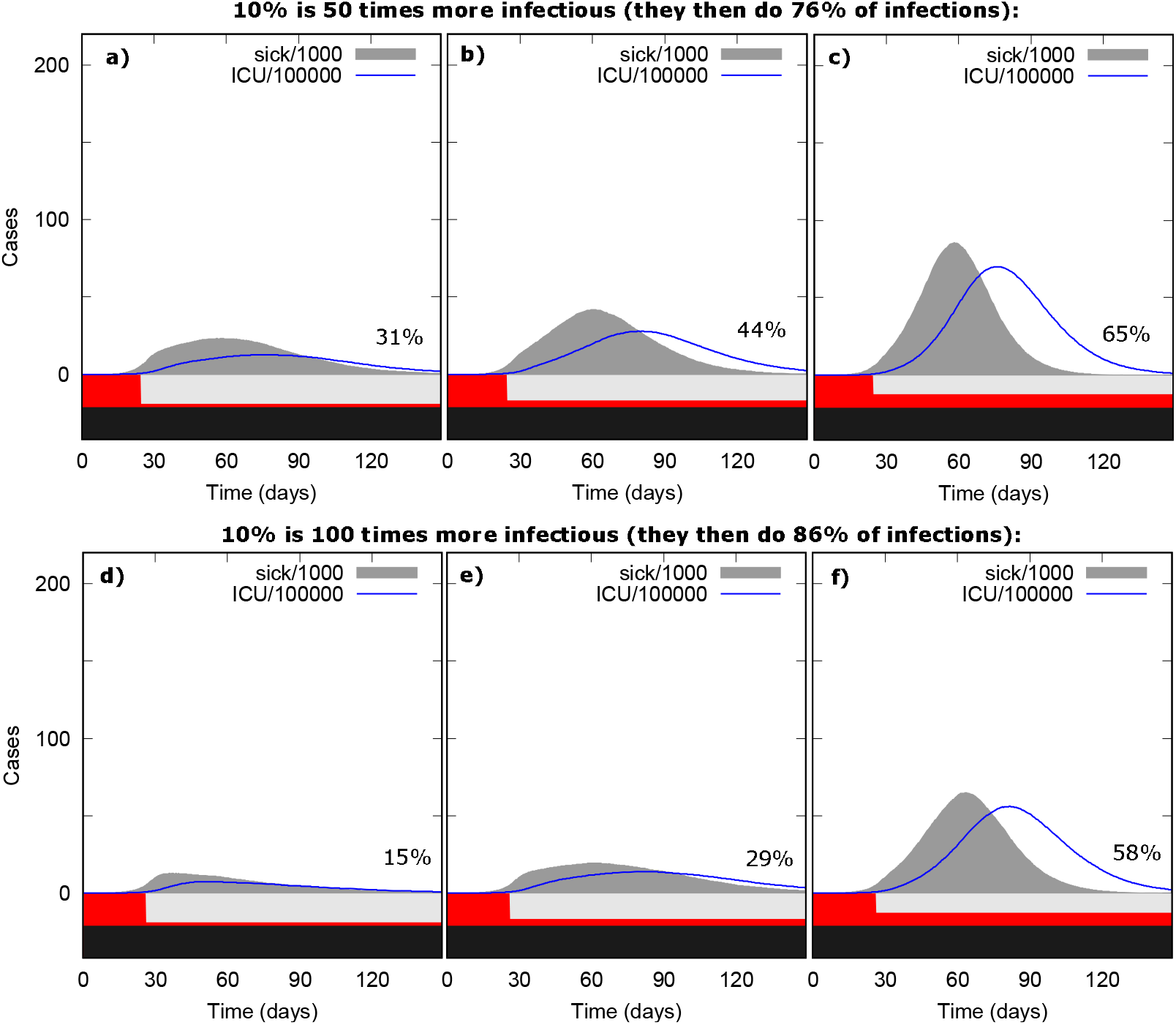
Sensitivity of results to opening “other” contacts. Panels a-c show the effect of increasing “other” contacts to 10%, 20% and 40% when s_i_ = 50; panels d-f show effect when s_i_ = 100. The black and red stacked bar shows the proportion of “work/school” and “other” contacts allowed.

**Figure S2.**
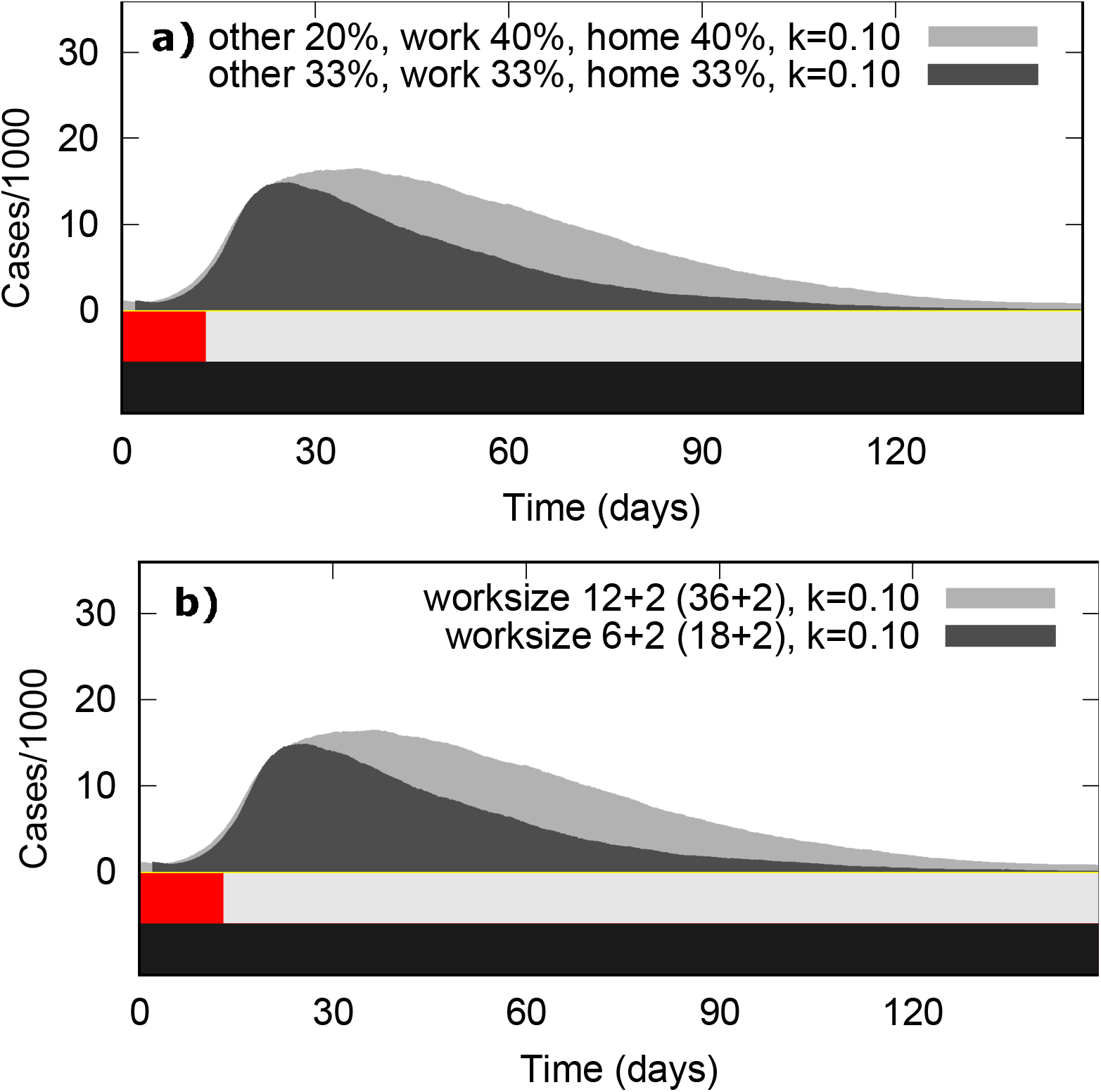
Sensitivity of results to proportion of contact type and size of workplace. The top panel (a) shows effect of reducing contact type “other” to 20% of total, while the bottom panel (b) shows effect of doubling the workplace to 12 members (with two more participating in a second workplace). The black and red stacked bar shows the proportion of “work/school” and “other” contacts allowed.

**Figure S3.**
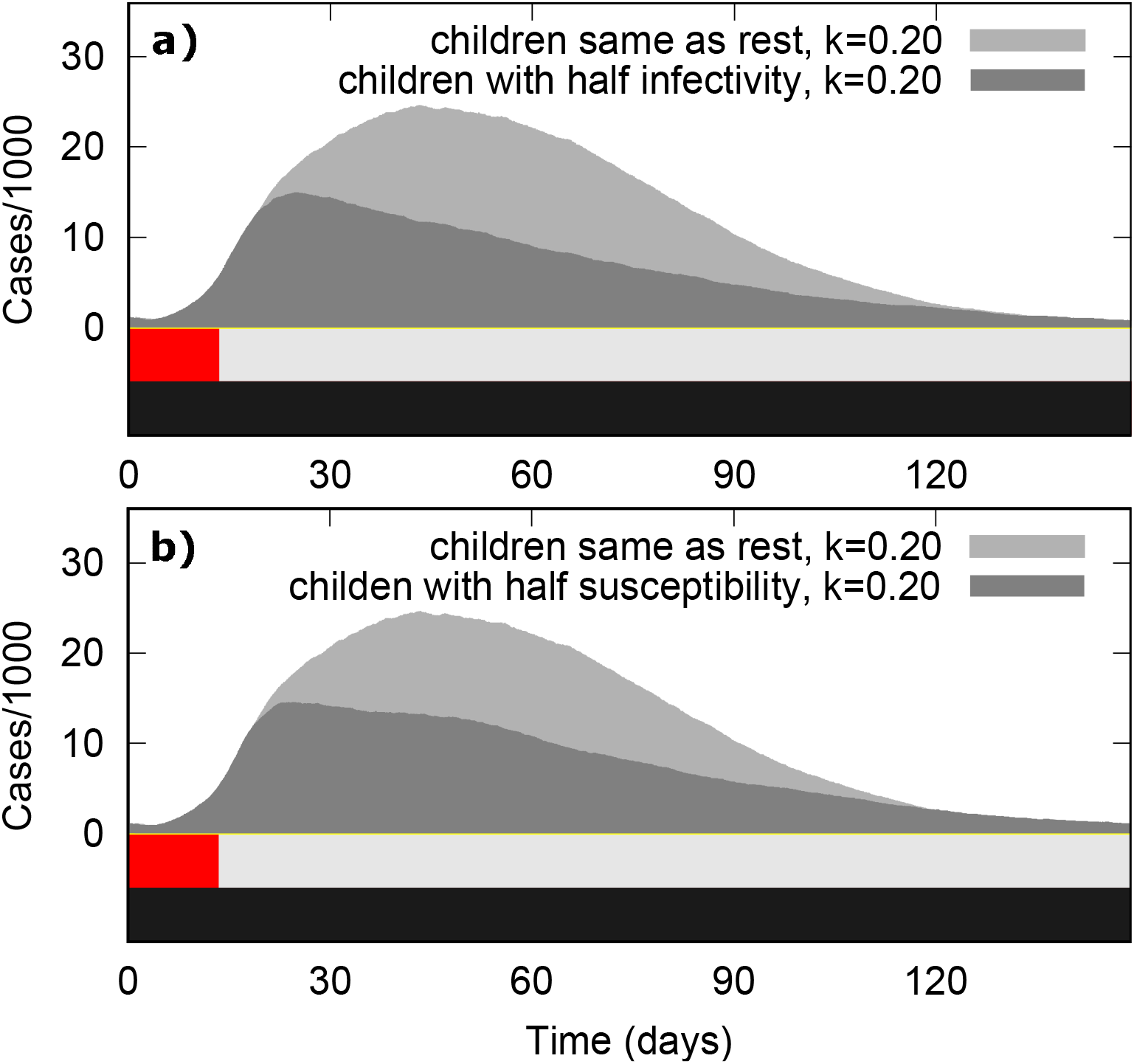
Sensitivity of results to children’s infectivity and susceptibility to infection. The top panel (a) shows effect of the effect of while the bottom panel (b) shows effect of doubling the workplace to 12 members (with two more participating in a second workplace). The black and red stacked bar shows the proportion of “work/school” and “other” contacts allowed.

**Table S1:** Distribution of simulated population by age groups, with conditional probabilities for hospitalization, Intensive Care Unit care or death,^1^ plus relative social contact.^2^ The hospitalization and ICU occupancy probabilities calibrated to an estimated infection fatality rate for COVID-19 of 0.3%.

| Age (years) | Percentage of population | Probability of hospitalization given infection | Probability of ICU given hospitalization | Probability of death given infection | Relative # social contacts |
| --- | --- | --- | --- | --- | --- |
| 0-9 | 10.9% | 0% | 5% | 0.029% | 1.21 |
| 10-19 | 11.9% | 0.013% | 5% | 0.029% | 1.70 |
| 20-29 | 13.3% | 0.37% | 5% | 0.020% | 1.45 |
| 30-39 | 11.7% | 1.1% | 5% | 0.029% | 1.45 |
| 40-49 | 13.6% | 1.4% | 6.3% | 0.059% | 1.38 |
| 50-59 | 13.6% | 2.7% | 12.2% | 0.137% | 1.31 |
| 60-69 | 11.7% | 3.9% | 27.4% | 0.450% | 1.06 |
| 70-79 | 8.9% | 5.5% | 43.2% | 1.133% | 0.81 |
| 80- | 4.3% | 5.5% | 70.9% | 2.06% | 0.81 |

## Notes

### Competing Interest Statement

The authors have declared no competing interest.

### Funding Statement

This project has received funding from the European Research Council (ERC) under the European Union's Horizon 2020 research and innovation program under grant agreement No. 740704, and from the Carlsberg Foundation in Denmark.

### Author Declarations

No IRB body involved here. It is a theoretical paper using public available data from Norwegian and Swedish health authorities.

